# Coffee Intake is Associated with Improved Insulin Sensitivity and Lower Visceral Adiposity: Evidence from Biomarker and Genetic Analysis

**DOI:** 10.64898/2026.06.25.26356610

**Authors:** Magdalena Sevilla-González, Xingyan Wang, Huan Yun, Zhendong Mei, Sarah Hsu, Paul A. Hanson, Jie Hu, Deirdre K. Tobias, Meryl S. LeBoff, Olga Demler, Aruna D. Pradhan, Samia Mora, I-Min Lee, Frank B. Hu, Miriam S. Udler, JoAnn E. Manson, Jun Li

## Abstract

**Importance:** Higher coffee intake has been associated with lower risk of type 2 diabetes (T2D), but the underlying biological pathways remain incompletely understood.

**Objective:** To examine associations of coffee intake with insulin sensitivity, adiposity, and T2D risk, and assess whether coffee intake modifies associations between pathway-specific genetic susceptibility and incident T2D.

**Design, Setting, and Participants:** Cross-sectional analyses among 806 participants without T2D in the VITamin D and OmegA-3 TriaL (VITAL) clinical sub-cohort, who underwent repeated dietary assessment, clinical phenotyping, and dual-energy X-ray absorptiometry imaging at baseline and year-2. Prospective analyses among 333,053 UK Biobank participants without T2D at baseline who had dietary and genetic data and were followed for a median of 13.3 years.

**Exposures:** Coffee intake assessed by food frequency questionnaires. In UK Biobank, 12 pathway-specific polygenic scores (pPS) representing distinct T2D pathophysiological mechanisms were evaluated.

**Main Outcomes and Measures:** The primary outcomes, in VITAL, were HbA1c, oral glucose tolerance test-derived measures of glucose response and insulin sensitivity, β-cell function, and overall, truncal, and visceral adiposity; in UK Biobank, was incident T2D.

**Results:** In VITAL, higher coffee intake was associated with higher insulin sensitivity (standardized β per cup/day, 0.046; P = .004) and lower visceral adipose tissue mass (β, −0.047; P = .006), after adjusting for demographic, lifestyle, and clinical factors, including body mass index. In UK Biobank, higher coffee intake was associated with lower T2D incidence (hazard ratio per cup/day, 0.96; 95% CI, 0.95-0.97), lower triglyceride-to-HDL cholesterol ratio (β,−0.01; P = 2.51 × 10^-19), and lower visceral adipose tissue mass (β, −0.01; P = 4.28 × 10^-9). Associations of 3 pPS related to insulin resistance and fat distribution with incident T2D were attenuated among participants consuming higher amount of coffee than among non-consumers (P for interaction < .0043).

**Conclusions and Relevance:** Higher coffee intake was associated with greater insulin sensitivity, lower visceral adiposity, and lower risk of T2D. Together with the attenuation of associations between pathway-specific genetic susceptibility and T2D risk among higher coffee consumers, these findings suggest that insulin resistance and visceral adiposity-related pathways may contribute to the association between coffee intake and T2D risk.

**Key Points:** *Question:* Is coffee intake associated with specific insulin sensitivity and adiposity markers, and type 2 diabetes risk, and does it modify associations between pathway-specific genetic susceptibility and type 2 diabetes?

*Findings:* In analyses repeated dietary, clinical, and imaging phenotyping in 806 VITAL participants and prospective data from 333,053 UK Biobank participants, higher coffee intake was associated with greater insulin sensitivity, lower visceral adiposity, and lower type 2 diabetes risk. Higher coffee intake also attenuated associations of three pathway-specific polygenic scores related to insulin resistance and fat distribution with type 2 diabetes risk.

*Meaning:* These findings suggest that pathways related to insulin sensitivity and visceral adiposity may contribute to the associations between coffee intake and lower type 2 diabetes risk.

## INTRODUCTION

Type 2 diabetes (T2D) is a chronic metabolic disorder characterized by high glycemia, insulin resistance, and pancreatic β-cell dysfunction^1^. It is one of the most prevalent non-communicable diseases globally, currently affecting 537 million people worldwide, with projections rising to 783 million by 2045^2^. T2D pathophysiology is complex and attributable to both genetic, environmental and lifestyle factors (e.g., diet).^3^ Recent research suggests that genetic predisposition to distinct pathophysiological pathways, including but not limited to impaired insulin secretion, insulin resistance, and ectopic fat deposition, contributes to T2D development.^4–9^

Suboptimal diets are estimated to account for approximately 70% of T2D cases globally.^10^ As one of the most widely consumed beverages worldwide, coffee has drawn significant attention for its potential health benefits.^11^ Roasted coffee contains ∼1,000 phytochemicals, including caffeine, chlorogenic acids (CGAs), trigonelline, and other polyphenols.^12^ Although caffeine intake has been shown to temporarily lower insulin sensitivity in acute feeding studies,^13^ over 30 large prospective cohort studies have consistently demonstrated a dose–response association between higher coffee consumption and reduced long-term risk of T2D.^14^ However, mechanisms by which coffee intake may be related to lower T2D risk remain largely unknown; elucidating these pathways may inform optimized dietary guidelines regarding coffee consumption for T2D prevention.

To address this knowledge gap, we investigated the relationship between habitual coffee intake and traits reflecting key T2D-related pathophysiological pathways, including glycemic burden, glucose response, insulin resistance, β-cell function, as well as overall, trunkal, and visceral adiposity, leveraging repeatedly phenotyping from the VITamin D and OmegA-3 TriaL (VITAL) clinical sub-cohort. Further, using process-specific polygenic risk scores (pPS)^4^ to reflect polygenic susceptibility for T2D through distinct pathophysiological pathways, we investigated whether coffee intake modified associations between pPS and long-term T2D risk, in the UK Biobank (UKBB) with 16 years of longitudinal follow-ups.

## METHODS

### Study Population

Details on VITAL and UK Biobank participants included in this study are provided in Supplementary Methods. Briefly, VITAL was a completed randomized, placebo-controlled trial of vitamin D3 and marine omega-3 fatty acid supplementation for the primary prevention of cancer and cardiovascular disease^15,16^. The VITAL-Clinical sub-cohort conducted detailed in-person phenotyping for 1,054 participants from the New England region, including questionnaires, interviews, oral glucose tolerance testing (OGTT), Dual-Energy X-Ray Absorptiometry (DXA) whole body composition scans, and biospecimen collection, at baseline and 2-year visit^17^. This analysis included 806 VITAL-Clinical sub-cohort participants who were free of T2D and had baseline dietary data and completed OGTT assessments at baseline and year 2. The Mass General Brigham Institutional Review Board approved the VITAL ancillary study, and all participants provided written informed consent. The UK Biobank is a prospective cohort of enrolling ∼500,000 adults aged 40-69 years during 2006-2010.^18^ This study included 333,053 participants who had coffee intake and genetic data available; were free of coronary heart disease, stroke, diabetes, and cancer; had HbA1c≤6.5% and did not report insulin use at baseline, and all analyses were conducted under application #27892.

### Assessment of coffee intake

In VITAL, coffee intake was assessed using a self-administered semiquantitative food frequency questionnaire at baseline and 4.5 years. Participants reported caffeinated and decaffeinated coffee separately, and responses were converted to servings per day. Year 2 coffee intake was estimated as the average of baseline and year 4.5 intake. Total coffee intake, and secondarily caffeinated and decaffeinated coffee intake, were modeled continuously in servings per day. In UK Biobank, information on the number of cups of coffee consumed per day and types of coffee regularly consumed was collected at baseline using a touchscreen questionnaire. Responses of less than 1 cup/day were coded as 0.5 cups/day, and values above 6 cups/day were winsorized to 6 cups/day to reduce the influence of extreme values. Additional data collection and coding details are provided in the Supplementary Methods.

### Assessment of blood biomarkers in VITAL-CTSC sub-cohort

We primarily selected four glycemic traits that reflected complementary pathways underlying T2D, including HbA1c (overall glycemic load), 2-hour glucose (post-load glucose metabolism) an insulin sensitivity index (ISI) from OGTT, and homeostasis model assessment of β-cell function (HOMA-B; β-cell function). We secondarily analyzed other fasting and post-load glucose and insulin measures, homeostatic model assessment for insulin resistance (HOMA-IR), metabolic clearance rate of glucose (MCRG), lipoprotein insulin resistance (LPIR), and diabetes risk index (DRI). We selected three primary adiposity traits to capture distinct aspects of body composition, including fat mass index (FMI), truncal fat mass, and visceral adipose tissue (VAT) mass. Body mass index (BMI), waist-to-hip ratio, and additional body composition traits including fat-to-lean mass ratio, were evaluated in secondary analyses. Detailed clinical biomarker assays^19^, trait definitions,^20,21^ equations,^22^ and DXA protocols^23^ are in the Supplementary Methods.

### UK Biobank outcomes and biomarkers

Incident T2D was defined using linked hospital inpatient records and death registry data based on ICD-10 codes E11.0-E11.9 occurring after baseline. Follow-up time was calculated from baseline assessment to T2D diagnosis, death, loss to follow-up, or the end of follow-up (May 31, 2022), whichever came first. Triglyceride-to-HDL cholesterol ratio, a surrogate biomarker for insulin resistance,^24^ and DXA-assessed VAT mass, were inverse-normal transformed before secondary analysis. Detailed outcome definitions are provided in the Supplementary Methods.

### UK Biobank pathway-specific polygenic scores and caffeine metabolism scores

Using recently published models, we calculated 12 pathway-specific polygenic scores (pPS) for T2D^4^ to capture genetic susceptibility through distinct pathophysiological pathways. Scores were standardized to mean=0 and standard deviation [SD]=1. Two additional weighted genetic scores were derived using previously identified caffeine-related variants (rs2472297, rs6968554, rs17685, and rs56113850) to account for genetic variation in caffeine metabolism.^25^ Details^28, 26,27^ are provided in the Supplementary Methods.

### Covariates

In VITAL, models were adjusted for age, sex, family history of diabetes, smoking status, race, alcohol use, physical activity, total juice intake, and red/processed meat intake; BMI was included in a separate model. In UK Biobank, models were adjusted for age, sex, smoking, alcohol intake, tea intake, physical activity, caffeine metabolism genetic scores, and in separate models, BMI. Analyses involving polygenic scores were additionally adjusted for genotyping batch, assessment center, and the top 10 genetic principal components. Detailed information on covariates is provided in the Supplementary Methods.

### Statistical analysis

In VITAL, skewed clinical and anthropometric traits were log-transformed as appropriate. Primary biomarkers were winsorized at +/-5 SD to reduce the influence of outliers and then standardized to mean=0 and SD=1. Associations between coffee intake and repeated glycemic and adiposity traits were estimated using linear mixed models with participant-specific random intercepts. Estimates represent SD differences in each trait per additional cup/day of coffee intake. Stratified analyses were further conducted by subgroups of age, BMI, physical activity, smoking status, sex, and family history of diabetes to evaluate potential effect modification. For the seven pre-specified primary traits, statistical significance was defined using a Bonferroni-corrected P <0.00714. Secondary traits were interpreted as exploratory, with P <0.05 considered nominally significant.

In UK Biobank, Cox regression was used to estimate hazard ratios (HR) and 95% confidence intervals (CI) for the association between coffee intake and incident T2D. Non-linear dose-response associations were evaluated using restricted cubic splines. We then examined associations between each pPS and T2D risk across categories of coffee intake and tested multiplicative interactions between coffee intake and each pPS using likelihood ratio tests. A Bonferroni-corrected P-interaction <0.00417 was used to define statistically interaction.

Complementary analyses of coffee intake with triglyceride-to-HDL cholesterol ratio and VAT mass were conducted using linear regression. All tests were two-sided. Additional analytic details are provided in the Supplementary Methods.

## RESULTS

### Coffee intake and T2D-related traits in VITAL Clinical sub-cohort

Our analysis included 806 VITAL participants (49.5% females, baseline median age: 65 years [Inter Quartile-Range (IQR): 60-69], BMI=27.2 kg/m^2^ [IQR: 24.4-30]) without T2D who had biomarker measurements available at baseline and year-2. Overall, 86% participants reported habitual coffee consumption. Median coffee intake was 1.79 cups/day (IQR, 0.79-2.50), primarily from caffeinated (median, 1 cups/day [IQR, 0.43-2.5]) rather than decaffeinated coffee (median, 0 cups/day [IQR, 0-0.07]) (**Table 1**).

**Table 1.** Basic characteristics of the 806 participants from the VITAL-CTSC.

| Characteristics | Values |
| --- | --- |
| **Age (years)** | 65 (60-69) |
| **Sex** |  |
| Female | 399 (50%) |
| Male | 407 (50%) |
| **BMI (kg/m<sup>2</sup>)** | 27.2 (24.4, 30.0) |
| **Primary biomarkers/traits** |  |
| **HbA1c (%)** | 5.30 (5.10-5.50) |
| **Glucose 120' (mg/dL)** | 115 (95-143) |
| **HOMA-B** | 75 (54-109) |
| **ISI** | 0.08 (0.06-0.10) |
| **FMI** | 9.1 (7.4-12) |
| **VAT mass** | 730 (498-962) |
| **Truncal Fat Mass** | 14,081 (10,483-17,730) |
| **Coffee intake (cups/day)** |  |
| **Caffeinated coffee** | 1.00 (0.43-2.50) |
| **Decaffeinated coffee** | 0.00 (0.00-0.07) |
| **Total Coffee** | 1.79 (0.79-2.50) |
| **Total Coffee Category** |  |
| 0 cup/d | 109 (14%) |
| ≤ 1 cup/d | 253 (31%) |
| 2–3 cups/d | 362 (45%) |
| 4+ cups/d | 82 (10%) |

Values are represented as median and interquartile range (IQR, 25th-75th), or n (%). VAT: Visceral Adipose Tissue, FMI: Fat Mass Index, ISI, Insulin Sensitivity Index.

Across primary glycemic traits, higher total coffee consumption was not associated with HbA1c (standardized β per cup/day =-0.022; *P* =0.33), 2-hr glucose (β =-0.026, *P* =0.22), or HOMA-B (β =-0.025, *P* =0.21), but was significantly associated with higher ISI (β =0.046, *P* =0.004) after adjusting for risk factors and BMI in linear mixed-effects analyses accounting for repeated measurements (**Figure 1**). In secondary biomarker analyses, higher coffee intake was associated with lower 2-hr insulin (β =-0.047, *P* =0.022) and higher MCRG (β =0.048, *P* =0.002), but not with fasting or 30min glucose or insulin, HOMA-IR, LPIR, or DRI (all *P*>0.05). These findings that higher coffee intake was more consistently related to whole-body insulin sensitivity than with fasting glycemia or β-cell function.

**Figure 1.**
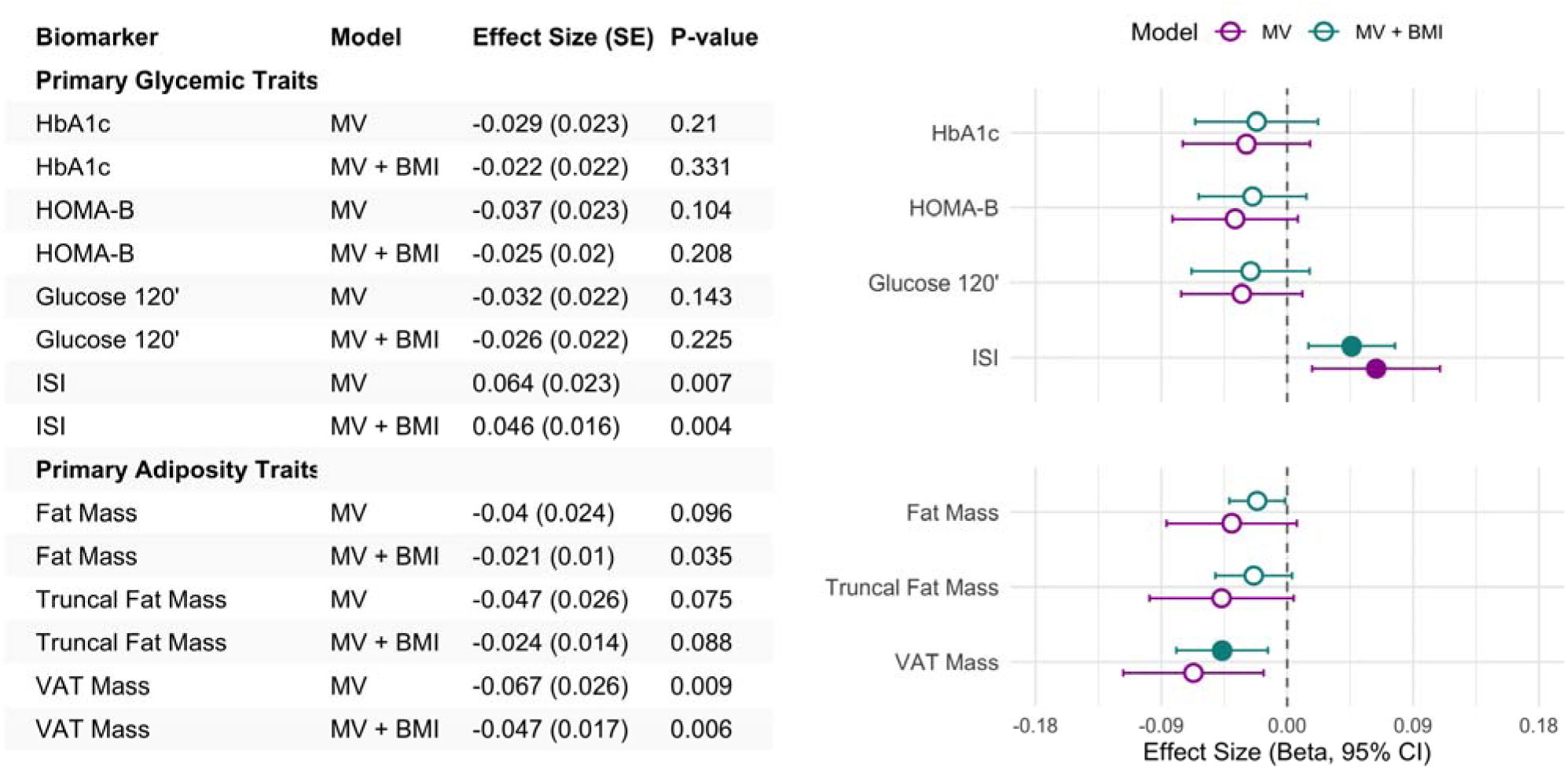
Association of total coffee intake and clinical biomarkers using multiple visits data in VITAL-CTSC sub-cohort. Data points represent the mean effect size (beta estimate). Error bars indicate 95% confidence intervals (CIs) calculated using linear mixed models per 1 standard deviation (SD). ISI Insulin Sensitivity Index, HOMA-B (Homeostasis Model Assessment Insulin Beta cell), VAT, Visceral Adipose Tissue. The following variables were log-transformed to improve normality: HOMA-B, 120-min glucose, Fat Mass Index (FMI), Trunk Fat Mass, VAT Mass. MV, Multivariable Model included covariates: age, biological sex, family history of diabetes, smoking status, self-report race, alcohol use, physical activity, juice, and red and processed meat consumption. Filled circles represent *p-value* < 0.0071 (Bonferroni adjustment)

For adiposity traits, higher total coffee consumption was associated with lower VAT mass in multivariable-adjusted analysis (β =-0.067, *P* =0.009); the association attenuated but remained significant after further adjusting for BMI (β =-0.047, *P* =0.006). In fully adjusted model, higher coffee intake was nominally associated with lower FMI (β =-0.021, *P* =0.035) but not truck fat mass (β =-0.024, *P* =0.09) (**Figure 1**). Among secondary adiposity traits, higher coffee intake was associated with lower VAT area (β =-0.046, *P* =0.005) and nominally associated with lower truck-to-limb fat percentage ratio (β =-0.046, *P* =0.022), lower fat mass–to–lean mass ratio (β =-0.028, *P* =0.019), but a modestly higher body weight (β =0.005, *P* =0.030).

All associations were generally consistent with or without BMI adjustment for BMI and appeared to be primarily driven by caffeinated coffee intake, although decaffeinated coffee intake was very low in this population (**Supplementary Table 1**). Associations were generally consistent across strata of age, sex, family history of T2D, and physical activity levels; however, inverse associations between coffee intake and FMI and VAT mass appeared to be stronger among females than males (*P-interaction* <0.05) (**Supplementary Figure 1**).

In secondary analyses, we did not observe significant associations between coffee intake and blood lipid measures (**Supplementary Table 1**).

### Coffee intake, insulin resistance, and visceral adiposity in UK Biobank

Among UK Biobank participants free of T2D at baseline, we first leveraged available biomarkers of insulin resistance (TG/HDL-C ratio, n=198,684) and visceral adiposity (VAT mass, n=21,424) to provide complementary evidence for the associations observed in VITAL. Higher coffee intake was associated with lower TG/HDL-C ratio (β =-0.010, *P* =2.5×10^-19^) and VAT mass (β =-0.015, *P* =4.28×10^-9^) in multivariable-adjusted models that included BMI and genetic risk scores related to caffeine metabolism. These associations were evident only after BMI adjustment for (**Supplementary Table 2**).

### Coffee intake, pPS, and T2D risk in UK Biobank

We next examined whether coffee consumption modified the associations between pPS and incident T2D among 333,053 UK Biobank participants free of T2D at baseline (**Table 2**). During a median follow-up of 13.3 years, 10,707 participants developed T2D.

**Table 2.** Baseline characteristics of study individuals in the UK Biobank.

| Characteristic | Overall,<br>N = 333,053<sup>1</sup> | No intake,<br>n = 73,795<sup>1</sup> | ≤1 cups/day<br>n = 90,463<sup>1</sup> | 2-3 cups/day<br>n = 103,183<sup>1</sup> | 4-5 cups/day<br>n = 44,601<sup>1</sup> | ≥6 cups/day<br>n = 20,314<sup>1</sup> | *P-value*<sup>2</sup> |
| --- | --- | --- | --- | --- | --- | --- | --- |
| Age (years) | 55 (8) | 54 (8) | 56 (8) | 56 (8) | 55 (8) | 54 (8) | <0.001 |
| Male, n (%) | 145,664 (44%) | 29,213 (40%) | 37,316 (41%) | 46,174 (45%) | 21,953 (49%) | 10,651 (52%) | <0.001 |
| **Ethnic Background** |  |  |  |  |  |  | <0.001 |
| Mixed | 7,528 (2.3%) | 2,631 (3.6%) | 2,317 (2.6%) | 1,769 (1.7%) | 538 (1.2%) | 224 (1.1%) |  |
| Black | 2,801 (0.8%) | 1,238 (1.7%) | 828 (0.9%) | 515 (0.5%) | 131 (0.3%) | 42 (0.2%) |  |
| Asian | 8,162 (2.5%) | 3,541 (4.8%) | 2,594 (2.9%) | 1,448 (1.4%) | 349 (0.8%) | 109 (0.5%) |  |
| White | 313,449 (94%) | 66,112 (90%) | 84,440 (93%) | 99,129 (96%) | 43,431 (97%) | 19,875 (98%) |  |
| Unknown/ Prefer<br>not to answer | 1,113 (0.3%) | 273 (0.4%) | 284 (0.3%) | 322 (0.3%) | 152 (0.3%) | 64 (0.3%) |  |
| BMI, kg/m<sup>2</sup> | 27.1 (4.6) | 27.1 (4.8) | 26.7 (4.5) | 27.0 (4.4) | 27.6 (4.6) | 28.0 (4.8) | <0.001 |
| HbA1c (%) | 5.34 (0.35) | 5.34 (0.36) | 5.34 (0.35) | 5.34 (0.34) | 5.34 (0.35) | 5.36 (0.36) | <0.001 |
| MET-minutes/week | 2,189 (2,674) | 2,238 (2,827) | 2,172 (2,581) | 2,176 (2,573) | 2,164 (2,685) | 2,268 (2,976) | <0.001 |
| **Smoking Status** |  |  |  |  |  |  | <0.001 |
| Current everyday | 24,632 (7.4%) | 5,193 (7.0%) | 4,216 (4.7%) | 6,275 (6.1%) | 4,555 (10%) | 4,284 (21%) |  |
| Current occasionally | 9,363 (2.8%) | 1,635 (2.2%) | 2,473 (2.7%) | 3,109 (3.0%) | 1,457 (3.3%) | 664 (3.3%) |  |
| Past | 191,789 (58%) | 21,272 (29%) | 28,991 (32%) | 34,654 (34%) | 15,450 (35%) | 6,755 (33%) |  |
| Never or unknown | 107,269 (32%) | 45,695 (62%) | 54,783 (61%) | 59,145 (57%) | 23,139 (52%) | 8,611 (42%) |  |

<sup>1</sup> Mean (SD); n (%)
<sup>2</sup> Kruskal-Wallis rank sum test; Pearson's Chi-squared test
BMI, body mass index; HbA1c, Hemoglobin A1c; MET, Metabolic Equivalent of Task.

Higher coffee intake was associated with lower T2D incidence, after adjusting for demographic factors and cardiometabolic risk factors, including BMI. Compared to non-coffee consumers, HR (95% CI) were 0.84 (0.79-0.88), 0.83 (0.79-0.88), 0.75 (0.70-0.81), and 0.83(0.77-0.91), for participants consuming ≤1, 2-3, 4-5, and ≥6 cups/day, respectively (*P-trend* <0.001). Associations were unchanged after further adjustment for genetic risk scores related to caffeine metabolism (**Table 3**). Restricted cubic spline analysis indicated a nonlinear dose-response relationship, with the strongest inverse association observed at 3-4 cups/day that gradually attenuated at higher intake levels (*P-linearity* <0.001, and *P-nonlinearity <0.001*) (**Supplementary Figure 2**).

**Table 3.** Associations Between Total Coffee Intake and Type 2 Diabetes risk in the UK Biobank.

|  | No intake | <=1 cups/day | 2-3 cups/day | 4-5 cups/day | >=6 cups/day | P-trend | HR (CI 95%)<br>per cup/day |
| --- | --- | --- | --- | --- | --- | --- | --- |
| **N** | 70,896 | 87,831 | 100,214 | 43,268 | 19,500 |  |  |
| **Cases** | 2,899 | 2,632 | 2,969 | 1,333 | 814 |  |  |
| **Person years** | 981,376 | 1,377,758 | 595,548 | 1,206,229 | 269,655 |  |  |
| **MV** | [Ref] | 0.81 (0.77-0.85) | 0.82 (0.77-0.86) | 0.79 (0.74-0.85) | 0.90 (0.83-0.97) | 1.24E-04 | 0.97 (0.96-0.98) |
| **MV + BMI** | [Ref] | 0.84 (0.79-0.88) | 0.83 (0.79-0.88) | 0.75 (0.70-0.81) | 0.83 (0.77-0.91) | 1.42E-09 | 0.96 (0.95-0.97) |
| **Further adjusted for caffeine CMS<sub>G2</sub>** |  |  |  |  |  |  |  |
| **MV** | [Ref] | 0.81 (0.77-0.85) | 0.82 (0.77-0.86) | 0.79 (0.74-0.85) | 0.90 (0.97-0.82) | 1.08E-04 | 0.97 (0.96-0.98) |
| **MV+BMI** | [Ref] | 0.84 (0.79-0.88) | 0.83 (0.79-0.88) | 0.76 (0.71-0.81) | 0.84 (0.77-0.91) | 2.23E-09 | 0.96 (0.95-0.97) |
| **Further adjusted for caffeine CMS<sub>G4</sub>** |  |  |  |  |  |  |  |
| **MV** | [Ref] | 0.81 (0.77-0.85) | 0.82 (0.77-0.86) | 0.79 (0.74-0.84) | 0.89 (0.82-0.97) | 8.89E-05 | 0.97 (0.96-0.98) |
| **MV+BMI** | [Ref] | 0.84 (0.79-0.88) | 0.83 (0.79-0.88) | 0.75 (0.70-0.81) | 0.84 (0.77-0.91) | 1.73E-09 | 0.96 (0.95-0.97) |

CMS<sub>G2</sub> included two SNPs: rs2472297 (near *CYP1A2*) and rs6968554 (near *AHR*). CMS<sub>G4</sub> included the two SNPs of CMS<sub>G2</sub> and included rs17685 (in *POR*) and rs56113850 (near *CYP2A6*). MV Cox Proportional Hazard model adjusted by biological sex, age, smoking frequency (“current everyday”, “current occasionally”, past and “never or unknow”) and quantity (“smoking pack years”), alcohol consumption (“daily or almost daily”, “three or four times a week”, “once or twice a week”, “one to three times a month”, “special occasions only”, “never”) , tea intake (cups/day), physical activity (MET minutes per week), and the top 10 genetic principal components. **Abbreviations:** MV Multivariable, BMI, Body Mass Index, CMS, Caffeine Metabolism Scores, CMS.

All 12 pPS representing different pathophysiological pathways were significantly associated with T2D after multivariable adjustment (**Supplementary Table 3**). Coffee consumption modified the associations between three pPS and T2D risk, including the alkaline phosphatase (ALP)-negative and liver-lipid genetic clusters, which remained significant after multiple-testing correction (multiplicative *P-interaction*<0.0041), and the SHBG–lipoprotein(a) metabolism cluster, which showed a nominally significant interaction (*P-interaction*=0.0044) (**Figure 2A and Supplementary Table 4**). These genetic clusters have previously been linked to lower corrected insulin response (CIR), HOMA-B, and gluteofemoral adipose tissue (GFAT), and higher proinsulin (PI) and VAT.^4^ Across the three pPS, genetic associations with T2D risk were the highest among non-coffee consumers, but progressively attenuated among participants with higher coffee intake (**Figure 2A**). No significant interactions were observed between coffee intake and the remaining pPS in relation to T2D risk (**Figure 2B**).

**Figure 2.**
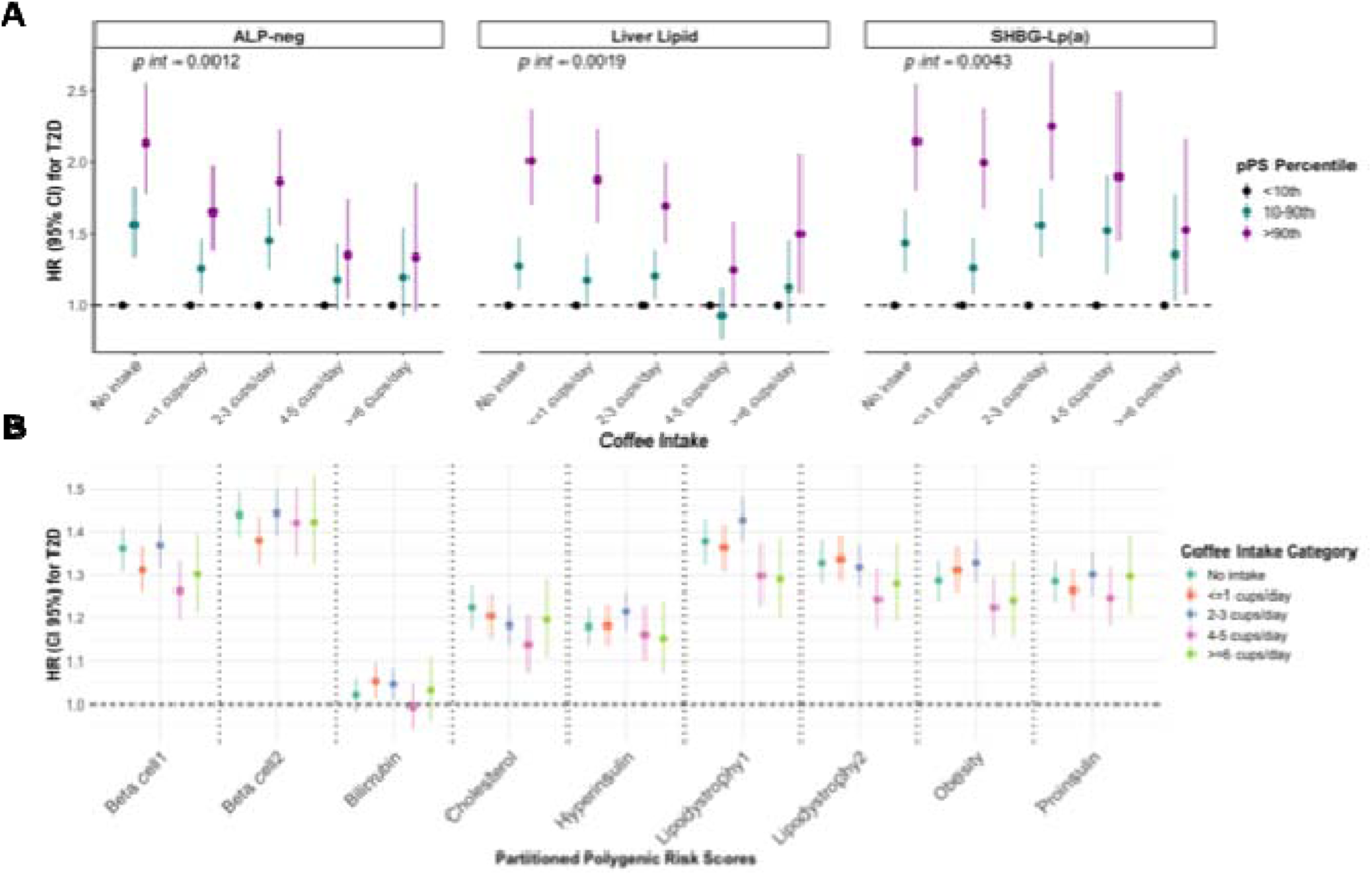
Association between coffee intake and partitioned polygenic risk scores (pPS) for type 2 diabetes (T2D). Data points represent hazard ratios (HRs) for incident T2D across categories of coffee intake, stratified by genetic risk group. Error bars indicate 95% confidence intervals (CIs), estimated using Cox proportional hazards models. Models were adjusted for sex, age, smoking frequency, smoking quantity, alcohol consumption, tea intake, physical activity, the top 10 genetic principal components, genotyping platform, and body mass index (BMI). Panel A included three pPS showing significant (*P-interaction <*0.0041) to nominal (*P-interaction <*0.05) interactions with total coffee intake in association with T2D risk based on the likelihood ratio tests; panel B shows results for other pPS.

## DISCUSSION

We investigated potential pathways underlying the association between higher coffee intake and lower T2D risk. In the VITAL-Clinical sub-cohort, higher coffee consumption was associated with enhanced insulin sensitivity and lower VAT mass, independent of established risk factors including BMI. In UK Biobank, we observed similar associations of coffee intake with lower insulin resistance and VAT mass. Additionally, the associations of 3 pathway-specific polygenic risk scores related to insulin resistance, insulin secretion, and fat distribution^4^ were attenuated among participants with higher coffee intake. These findings suggest that improved insulin sensitivity and reduced visceral adiposity may contribute to the associations between higher coffee consumption and lower T2D risk.

Although coffee or caffeine ingestion may transiently impair insulin sensitivity,^28–30, 31^ epidemiologic studies consistently associate habitual coffee consumption with improved insulin sensitivity^32–35^ and lower long-term T2D risk.^36^ Our findings extend this evidence by linking coffee intake to comprehensive OGTT-derived measures of glucose metabolism. In VITAL, higher coffee intake was associated with higher ISI and MCRG, and lower 2-hr insulin, but not with fasting glycemia, HOMA-B, or 2-hr glucose. These findings suggest that coffee intake may be more strongly related to whole-body insulin sensitivity and postprandial glucose disposal than fasting glycemia or insulin secretion. Our results are consistent with those from the Swedish Uppsala Longitudinal Study of Adult Men, in which coffee consumption was associated with greater insulin sensitivity measured by a hyper insulinemic-euglycemic clamp after adjustment for correlated dietary/lifestyle factors.^37^ Similarly, we observe no association between coffee intake and markers of early insulin response/insulin secretion, including 30-min glucose and insulin levels. Although a recent meta-analysis of clinical trials reported that coffee intake improved HOMA-IR (*P*=0.01) but not the Matsuda ISI (*P*=0.08), included studies were generally small and heterogeneous.^38^

Our genetic interaction analysis in UK Biobank further supported a role of insulin resistance in coffee-T2D risk associations. Of the pPS whose association with T2D risk were attenuated by coffee consumption, the liver-lipid cluster is primarily driven by variants mapped to *GCKR* and *FADS1* (involved in lipid metabolism)^4^ and the ALP-negative cluster included variants on *ABO*, *FADS1, TM6SF2* (regulates hepatic fat metabolism)*, and GCKR. GCKR* encodes a protein that regulates hepatic glucokinase (GCK), which converts glucose to glucose-6-phosphate (G6P), a critical in hepatic glucose uptake, glucose utilization, and post-load glucose clearance.^39^ Interestingly, CGAs, a class of phenolic compounds abundant in coffee, may reduce hepatic glucose output by inhibiting glucose-6-phosphatase, an opposing enzyme of GCK that converts G6P into glucose.^40^ Other studies suggests that CGAs may regulate GLUT4, thus improving post-load glucose metabolism.^41^ Both the liver-lipid and ALP-negative clusters were previously linked to lower CIR and HOMA-IR.^4^ Together with our biomarker results, these findings suggests insulin sensitivity and postprandial glucose clearance as a potential pathway linking coffee intake with lower T2D risk.

In addition to glycemic traits, higher coffee intake was associated with lower VAT independent of BMI in both VITAL-Clinical sub-cohort and UK Biobank. In a study among Japanese adults, higher coffee consumption was associated with lower visceral obesity,^42^ and in PREDIMED-PLUS, increasing caffeinated coffee consumption from low to moderate was associated with reduced total body fat, trunk fat, and VAT;^43^ neither study adjusted for BMI in multivariable models. Notably in VITAL, coffee intake was not associated with BMI, WHR, or total fat mass, and its association with VAT persisted after BMI adjustment; the inverse association of coffee with VAT in UK Biobank also became evident after adjusting for BMI. These findings extend prior evidence, suggesting that coffee maybe more closely related to fat distribution rather than overall obesity alone. In a 12-week RCT, high-CGA *vs.* low-CGA coffee interventions led to reductions VAT,^44^ supporting CGA as a potential agent underlying the observed associations. Nonetheless, whether these associations are attributable to caffeine^45^ or other coffee phytochemicals remain uncertain.

Our genetic interaction analysis further supported these DXA findings. The ALP-negative and SHBG-lipoprotein(a) genetic clusters have been previously linked to higher BMI-adjusted VAT and lower GFAT, respectively.^4^ VAT accumulation is thought to be a major pathway by which obesity increases T2D risk and has been associated with T2D risk independently of BMI.^46^ Because VAT is drained by the portal vein, excessive VAT may expose the liver to high levels of free fatty acids and glycerol, contributing to hepatic insulin resistance and increased hepatic glucose production.^47^ In contrast, GFAT has been linked to favorable cardiometabolic profiles^48^. Variants in *SHBG* are genetic determinants for GFAT,^49^ and higher circulating sex hormone–binding globulin levels have been associated with lower T2D risk.^50^ Our findings and previous evidence collectively suggested that body fat distribution, particularly lower VAT, may be a potential pathway linking coffee intake to lower T2D risk.

The strengths of our study include the corroborative evidence from biomarker analysis and genetic analysis from two independent, well-characterized cohorts. Comprehensive OGTT-derived glycemic measures, DXA-derived body composition measures, and pathway-specific pPS allowed us to examine potential mechanisms linking coffee consumption to T2D risk.

Several limitations should be noted. First, the observational design precludes causal inference. Although we adjusted for key confounders, residual confounding from unmeasured lifestyle or dietary factors cannot be excluded. Second, self-reported coffee intake may be subject to bias in recall and estimation, with limited information on preparation methods or additives. Third, we have limited ability to compare between caffeinated and decaffeinated coffee given the very low consumption decaffeinated coffee in VITAL and limited beverage questionnaire in UK Biobank. Finally, although we analyzed two independent cohorts, the majority of our participants were non-Hispanic White middle-aged to older adults, which may limit the generalizability of our findings other populations.

In conclusion, higher coffee intake was associated with biomarkers of greater insulin sensitivity and lower VAT mass. Genetic analyses further showed that coffee intake was associated with attenuated polygenic risk for T2D through pathways relevant to insulin resistance and fat distribution. Future studies, especially randomized controlled trials that compare caffeinated and decaffeinated coffee and non-coffee control beverages, are warranted to determine causality, clarify mechanisms underlying these associations, and distinguish the effects of caffeine from those of other bioactive coffee constituents, therefore to better inform dietary guidelines for coffee consumption.

## DISCLOSURES

MS-G and MSU are involved in research funded in collaboration with Novo Nordisk. MSU also has a consulting activity with Novo Nordisk.

## DATA SHARING STATEMENT

VITAL data are available from the study investigators upon reasonable request and appropriate approval. UK Biobank data are available to approved researchers through the UK Biobank Research Analysis Platform (https://www.ukbiobank.ac.uk). Analytic code may be available from the corresponding author upon reasonable request.

## FUNDING

MS-G is supported by the NIH NIDDK grant and 1K99DK139461-01A1 and 5U24DK132733-02. MSU is supported by the Doris Duke Foundation (Award 2022063). The Vitamin D and Omega-3 Trial was supported by grant R01AT011729, R01DK112940, R01HL134811, R01HL160799, K24HL136852 from the National Center for Complementary and Integrative Health and, during the intervention phase, was supported by grants U01 CA138962 and R01 CA138962 from the National Cancer Institute; National Heart, Lung, and Blood Institute; Office of Dietary Supplements; National Institute of Neurological Disorders and Stroke; and the National Center for Complementary and Integrative Health. The ancillary studies are supported by grants from multiple institutes, including the National Heart, Lung, and Blood Institute; the National Institute of Diabetes and Digestive and Kidney Diseases; the National Institute on Aging; the National Institute of Arthritis and Musculoskeletal and Skin Diseases; the National Institute of Mental Health; and others. Pharmavite LLC of Northridge, California (vitamin D) and Pronova BioPharma of Norway and BASF (Omacor fish oil) donated the study agents, matching placebos, and packaging in the form of calendar packs. Quest Diagnostics and Labcorp performed the VITAL biomarker laboratory measurements at no additional costs to the VITAL study. The funding sources had no role in the design and conduct of this study or the interpretation of the data. The opinions expressed in the manuscript are those of the study authors.

## Data Availability

All data produced in the present study are available upon reasonable request to the authors

## Notes

### Author Declarations

The Mass General Brigham Institutional Review Board approved this ancillary study, and all participants provided written informed consent before enrollment in VITAL.

## References

1. Lu, X. et al. Type 2 diabetes mellitus in adults: pathogenesis, prevention and therapy. Signal Transduction and Targeted Therapy 2024 9:1 9, 1–25 (2024).

2. What Is Diabetes | International Federation of Diabetes. https://idf.org/about-diabetes/what-is-diabetes/.

3. Nayor, M., Brown, K. J. & Vasan, R. S. The Molecular Basis of Predicting Atherosclerotic Cardiovascular Disease Risk. Circ. Res. 128, 287–303 (2021).

4. Smith, K. et al. Multi-ancestry polygenic mechanisms of type 2 diabetes. Nat. Med. https://doi.org/10.1038/S41591-024-02865-3 (2024) doi:10.1038/S41591-024-02865-3.

5. Kim, H. et al. High-throughput genetic clustering of type 2 diabetes loci reveals heterogeneous mechanistic pathways of metabolic disease. Diabetologia 2022 66:3 66, 495–507 (2022).

6. Suzuki, K. et al. Genetic drivers of heterogeneity in type 2 diabetes pathophysiology. Nature 627, 347–357 (2024).

7. DiCorpo, D. et al. Type 2 Diabetes Partitioned Polygenic Scores Associate With Disease Outcomes in 454,193 Individuals Across 13 Cohorts. Diabetes Care 45, 674–683 (2022).

8. Udler, M. S. et al. Type 2 diabetes genetic loci informed by multi-trait associations point to disease mechanisms and subtypes: A soft clustering analysis. PLoS Med. 15, e1002654 (2018).

9. Sevilla-González, M. et al. Heterogeneous effects of genetic variants and traits associated with fasting insulin on cardiometabolic outcomes. Nat. Commun. 16, 2569 (2025).

10. O’Hearn, M. et al. Incident type 2 diabetes attributable to suboptimal diet in 184 countries. Nat. Med. 29, 982–995 (2023).

11. van Dam, R. M., Hu, F. B. & Willett, W. C. Coffee, Caffeine, and Health. N. Engl. J. Med. 383, 369–378 (2020).

12. Loftfield, E. et al. Association of coffee drinking with mortality by genetic variation in caffeine metabolism: Findings from the UK Biobank. JAMA Intern. Med. 178, 1086–1097 (2018).

13. Keijzers, G. B., De Galan, B. E., Tack, C. J. & Smits, P. Caffeine can decrease insulin sensitivity in humans. Diabetes Care 25, 364–369 (2002).

14. Ding, M., Bhupathiraju, S. N., Chen, M., Van Dam, R. M. & Hu, F. B. Caffeinated and decaffeinated coffee consumption and risk of type 2 diabetes: a systematic review and a dose-response meta-analysis. Diabetes Care 37, 569–586 (2014).

15. Manson, J. E. et al. Marine n-3 Fatty Acids and Prevention of Cardiovascular Disease and Cancer. N. Engl. J. Med. 380, 23–32 (2019).

16. Manson, J. E. et al. Vitamin D Supplements and Prevention of Cancer and Cardiovascular Disease. N. Engl. J. Med. 380, 33–44 (2019).

17. Orkaby, A. R. et al. Effect of Vitamin D3 and Omega-3 Fatty Acid Supplementation on Risk of Frailty: An Ancillary Study of a Randomized Clinical Trial. *JAMA Netw*. Open 5, E2231206 (2022).

18. Sudlow, C. et al. UK biobank: an open access resource for identifying the causes of a wide range of complex diseases of middle and old age. PLoS Med. 12, (2015).

19. Qian, F. et al. Biomarkers of glucose-insulin homeostasis and incident type 2 diabetes and cardiovascular disease: results from the Vitamin D and Omega-3 trial. Cardiovasc. Diabetol. 23, (2024).

20. Estimating Visceral Fat By Dual-Energy X-Ray Absorptiometry. (2010).

21. Bea, J. W. et al. MRI Based Validation of Abdominal Adipose Tissue Measurements From DXA in Postmenopausal Women. Journal of Clinical Densitometry 25, 189–197 (2022).

22. Stumvoll, M., Van Haeften, T., Fritsche, A. & Gerich, J. Oral glucose tolerance test indexes for insulin sensitivity and secretion based on various availabilities of sampling times. Diabetes Care 24, 796–797 (2001).

23. Neeland, I. J. et al. Visceral and ectopic fat, atherosclerosis, and cardiometabolic disease: a position statement. Lancet Diabetes Endocrinol. 7, 715–725 (2019).

24. McLaughlin, T. et al. Is there a simple way to identify insulin-resistant individuals at increased risk of cardiovascular disease? American Journal of Cardiology 96, 399–404 (2005).

25. Cornelis, M. C. et al. Genome-wide association study of caffeine metabolites provides new insights to caffeine metabolism and dietary caffeine-consumption behavior. Hum. Mol. Genet. 25, 5472–5482 (2016).

26. Bycroft, C. et al. The UK Biobank resource with deep phenotyping and genomic data. Nature 562, 203–209 (2018).

27. Loftfield, E. et al. Association of Coffee Drinking With Mortality by Genetic Variation in Caffeine Metabolism: Findings From the UK Biobank. JAMA Intern. Med. 178, 1086–1097 (2018).

28. Lane, J. D., Barkauskas, C. E., Surwit, R. S. & Feinglos, M. N. Caffeine impairs glucose metabolism in type 2 diabetes. Diabetes Care 27, 2047–2048 (2004).

29. Petrie, H. J. et al. Caffeine ingestion increases the insulin response to an oral-glucose-tolerance test in obese men before and after weight loss. American Journal of Clinical Nutrition 80, 22–28 (2004).

30. Jagim, A. R. et al. International society of sports nutrition position stand: energy drinks and energy shots. J. Int. Soc. Sports Nutr. 20, (2023).

31. Keijzers, G. B., De Galan, B. E., Tack, C. J. & Smits, P. Caffeine can decrease insulin sensitivity in humans. Diabetes Care 25, 364–369 (2002).

32. Yamaji, T. et al. Coffee consumption and glucose tolerance status in middle-aged Japanese men. Diabetologia 47, 2145–2151 (2004).

33. Smith, B., Wingard, D. L., Smith, T. C., Kritz-Silverstein, D. & Barrett-Connor, E. Does coffee consumption reduce the risk of type 2 diabetes in individuals with impaired glucose? Diabetes Care 29, 2385–2390 (2006).

34. Wu, T., Willett, W. C., Hankinson, S. E. & Giovannucci, E. Caffeinated coffee, decaffeinated coffee, and caffeine in relation to plasma C-peptide levels, a marker of insulin secretion, in U.S. women. Diabetes Care 28, 1390–1396 (2005).

35. Ärnlöv, J., Vessby, B. & Risérus, U. Coffee Consumption and Insulin Sensitivity [4]. JAMA 291, 1199–1201 (2004).

36. Ding, M., Bhupathiraju, S. N., Chen, M., Van Dam, R. M. & Hu, F. B. Caffeinated and decaffeinated coffee consumption and risk of type 2 diabetes: a systematic review and a dose-response meta-analysis. Diabetes Care 37, 569–586 (2014).

37. Ärnlöv, J., Vessby, B. & Risérus, U. Coffee Consumption and Insulin Sensitivity [4]. JAMA 291, 1199–1201 (2004).

38. Moon, S. M., Joo, M. J., Lee, Y. S. & Kim, M. G. Effects of coffee consumption on insulin resistance and sensitivity: A meta-analysis. Nutrients 13, (2021).

39. Abu Aqel, Y. et al. Glucokinase (GCK) in diabetes: from molecular mechanisms to disease pathogenesis. Cell. Mol. Biol. Lett. 29, 1–27 (2024).

40. Kumar Ramalingam, P. et al. In Silico Screening of Chlorogenic Acids from Plant Sources against Human Translocase-I to Identify Competitive Inhibitors to Treat Diabetes. ACS Omega 9, 6561–6568 (2024).

41. Meng, S., Cao, J., Feng, Q., Peng, J. & Hu, Y. Roles of Chlorogenic Acid on Regulating Glucose and Lipids Metabolism: A Review. Evid. Based. Complement. Alternat. Med. 2013, 801457 (2013).

42. Koyama, T., Maekawa, M., Ozaki, E., Kuriyama, N. & Uehara, R. Daily consumption of coffee and eating bread at breakfast time is associated with lower visceral adipose tissue and with lower prevalence of both visceral obesity and metabolic syndrome in Japanese populations: A cross-sectional study. Nutrients 12, 1–10 (2020).

43. Henn, M. et al. Increase from low to moderate, but not high, caffeinated coffee consumption is associated with favorable changes in body fat. Clinical Nutrition 42, 477–485 (2023).

44. Watanabe, T. et al. Coffee abundant in chlorogenic acids reduces abdominal fat in overweight adults: A randomized, double-blind, controlled trial. Nutrients 11, (2019).

45. van Dam, R. M., Hu, F. B. & Willett, W. C. Coffee, Caffeine, and Health. New England Journal of Medicine 383, 369–378 (2020).

46. Agrawal, S. et al. BMI-adjusted adipose tissue volumes exhibit depot-specific and divergent associations with cardiometabolic diseases. Nat. Commun. 14, (2023).

47. Neeland, I. J. et al. Visceral and ectopic fat, atherosclerosis, and cardiometabolic disease: a position statement. Lancet Diabetes Endocrinol. 7, 715–725 (2019).

48. Agrawal, S. et al. Inherited basis of visceral, abdominal subcutaneous and gluteofemoral fat depots. Nat. Commun. 13, 1–17 (2022).

49. Manolopoulos, K. N., Karpe, F. & Frayn, K. N. Gluteofemoral body fat as a determinant of metabolic health. Int. J. Obes. 34, 949–959 (2010).

50. Ding, E. L. et al. Sex Hormone–Binding Globulin and Risk of Type 2 Diabetes in Women and Men. New England Journal of Medicine 361, 1152–1163 (2009).

