## Supplementary Figures for "Coffee Intake is Associated with Improved Insulin Sensitivity and Lower Visceral Adiposity: Evidence from Biomarker and Genetic Analysis"

**Supplementary Figure 1. Stratified analysis by clinical risk factors and total coffee intake in VITAL-CTSC sub-cohort.** Data points represent the mean effect size (beta estimate). Error bars indicate 95% confidence intervals (CIs) calculated using linear mixed models. ISI Insulin Sensitivity Index, FMI, Fat Mass Index, VAT, Visceral Adipose Tissue. MV: Multivariable Model included covariates: age, biological sex, family history of diabetes, smoking status, self-report race, alcohol use, physical activity, juice, and red and processed meat consumption. FMI and VAT mass were log-transformed to improve normality.

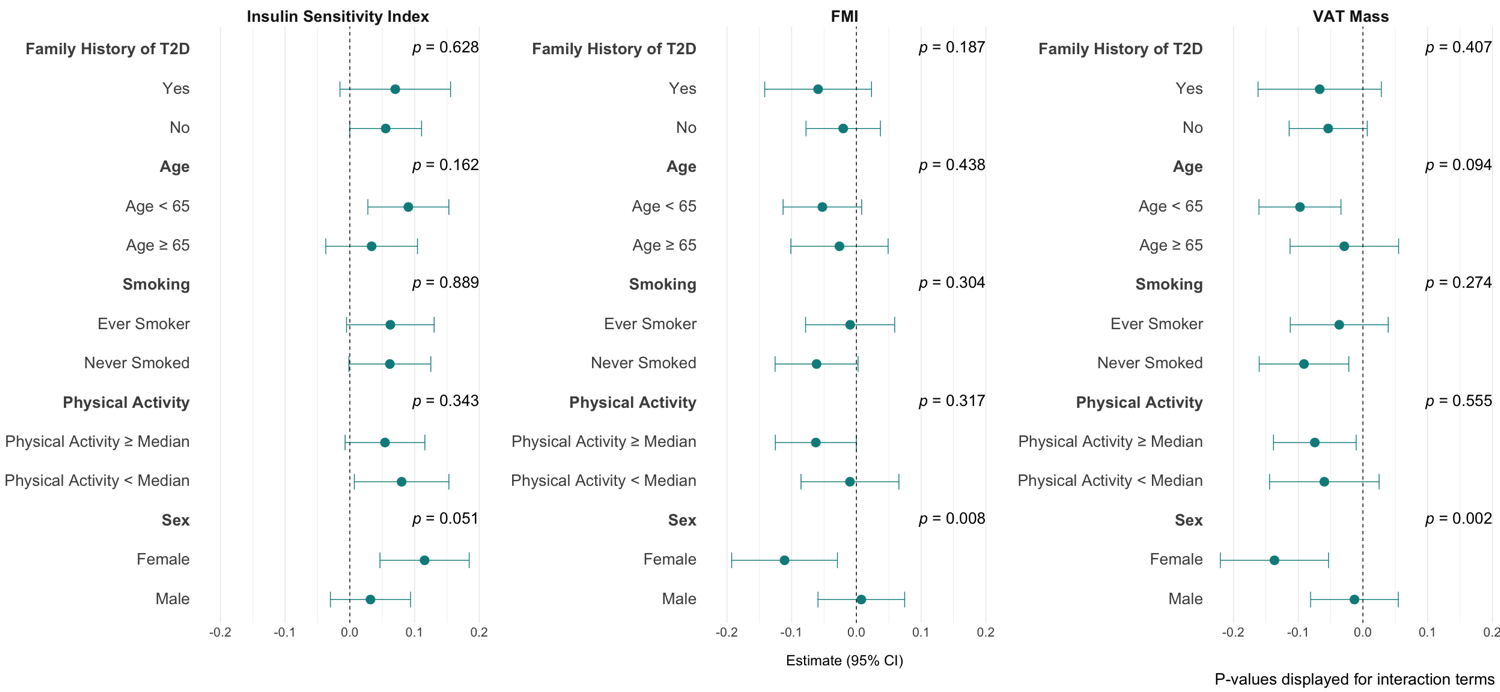

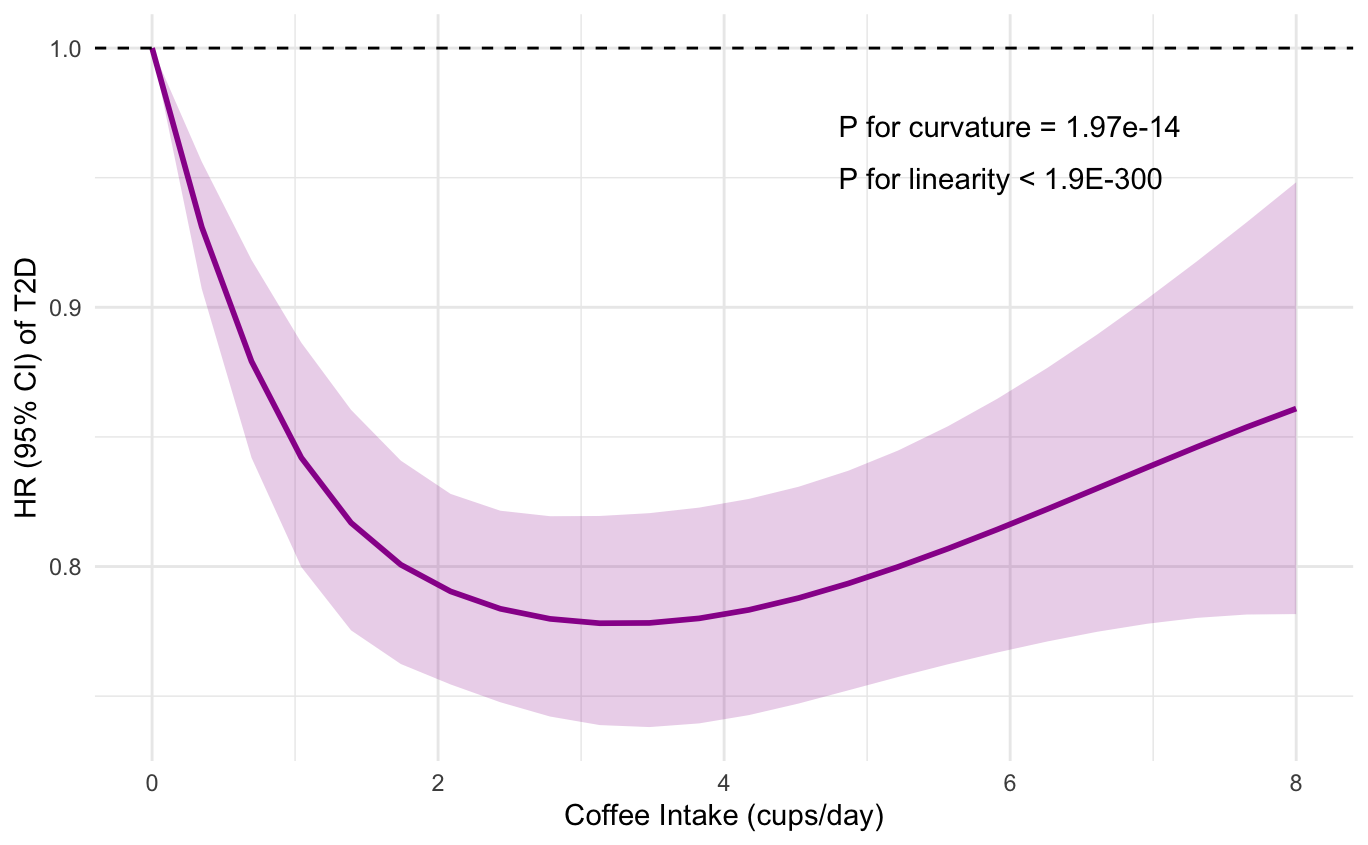
**Supplementary Figure 2. Dose-response relationship of coffee intake with risk of type 2 diabetes (T2D) in the UKBB** estimated using a restricted cubic spline model. The solid purple line represents the hazard ratio (HR), and the shaded area indicates the 95% confidence interval (CI). The dashed horizontal line denotes the reference HR of 1.0. P values for linearity and curvature were estimated using Wald tests of the spline.
