## Supplementary Methods for "Coffee Intake is Associated with Improved Insulin Sensitivity and Lower Visceral Adiposity: Evidence from Biomarker and Genetic Analysis"

**VITAL CTSC sub-cohort study population**

The VITamin D and OmegA-3 TriaL (VITAL; NCT01169259) was a completed randomized, placebo-controlled, 2 x 2 factorial, double-blind clinical trial evaluating daily vitamin D3 supplementation at 2000 IU/day and marine omega-3 fatty acid supplementation at 1 g/day for the primary prevention of cancer and cardiovascular disease. The trial enrolled 25,871 U.S. adults, including men aged >=50 years and women aged >=55 years.

A sub-cohort of 1,054 VITAL participants from the New England region was enrolled in detailed in-person clinical phenotyping at the Brigham and Women's Hospital Harvard Catalyst Clinical and Translational Science Center in Boston, Massachusetts. Participants completed visits at baseline and at the 2-year follow-up. At each visit, participants were interviewed regarding demographic characteristics, medical history, and lifestyle factors; completed dietary questionnaires; provided fasting blood samples; underwent a 2-hour oral glucose tolerance test; and completed dual-energy X-ray absorptiometry-based body composition assessment.

For the present analysis of coffee intake and type 2 diabetes-related glycemic and adiposity traits, we excluded participants with prevalent type 2 diabetes at enrollment, those who did not complete the oral glucose tolerance test at both baseline and year 2, and those with missing baseline dietary data. The final analytic sample included 806 participants. The Mass General Brigham Institutional Review Board approved this ancillary study, and all participants provided written informed consent before enrollment in VITAL.

### UK Biobank study population

The UK Biobank is a population-based prospective cohort that enrolled approximately 500,000 participants aged 40-69 years between 2006 and 2010 through the United Kingdom's National Health Service. At baseline, participants completed touchscreen questionnaires, underwent physical measurements, and provided biological samples. Genetic data were available for a large subset of participants.

For the analysis of coffee intake, pathway-specific polygenic scores, and incident type 2 diabetes, we included participants with baseline coffee intake and genetic data. Participants were excluded if they reported a history of coronary heart disease, stroke, type 1 or type 2 diabetes, or cancer before baseline. Prevalent coronary heart disease, stroke, and diabetes were identified using UK Biobank self-reported disease fields 6150 and 2443 at instance 0. Cancer diagnoses before baseline were identified using cancer diagnosis date field 40005. We additionally excluded participants with baseline HbA1c >6.5% or self-reported insulin use at baseline, assessed using field 6177 for men and field 6153 for women at instance 0. The final analytic sample included 333,053 participants. UK Biobank analyses were conducted under application #27892.

**Assessment and coding of coffee intake**

In VITAL, coffee intake was assessed using a self-administered semiquantitative food frequency questionnaire administered at baseline and 4.5 years after baseline. The questionnaire assessed usual intake of approximately 60 foods and beverages over the previous year and was adapted to prioritize dietary sources of vitamin D and omega-3 fatty acids.

Participants reported caffeinated and decaffeinated coffee intake separately. One serving was defined as one 8 oz cup. Frequency categories were: never, 1-3 times/month, 1 time/week, 2-4 times/week, 5-6 times/week, 1 time/day, 2-3 times/day, 4-5 times/day, and >=6 times/day. These categories were converted to estimated servings per day by assigning an average daily intake to each category. Total coffee intake was calculated as the sum of caffeinated and decaffeinated coffee intake. Under the assumption that coffee intake remained generally stable over time with modest gradual changes, estimated coffee intake at year 2 was calculated as the average of baseline and year 4.5 intake. Baseline and estimated year 2 total coffee intake, and secondarily caffeinated and decaffeinated coffee intake, were modeled as continuous variables in servings per day.

In UK Biobank, total coffee intake was assessed at baseline using touchscreen questionnaire field 1498, instance 0, which asked: 'How many cups of coffee do you drink each day? Include decaffeinated coffee.' Participants could report a numeric number of cups per day or select 'Less than 1,' 'Do not know,' or 'Prefer not to answer.' Participants selecting 'Do not know' or 'Prefer not to answer' were treated as missing for coffee intake. Responses of 'Less than 1' were coded as 0.5 cups/day. To reduce the influence of extreme values and potential reporting error, coffee intake was winsorized at the 90th percentile; values greater than 6 cups/day were assigned a value of 6 cups/day.

For categorical analyses, coffee intake was categorized as none, <=1 cup/day, 2-3 cups/day, 4-5 cups/day, and >=6 cups/day, with no coffee intake used as the reference category.

**Blood biomarker measurements in VITAL CTSC**

Blood samples were collected during the oral glucose tolerance test at fasting, 30 minutes, and 2 hours at both the baseline and year 2 visits. Samples from baseline and follow-up were paired and assayed in random order to minimize batch effects.

Plasma glucose and insulin concentrations were measured using enzymatic assays on the Roche Cobas 6000 system with Roche Diagnostics reagents. HbA1c was measured using the Roche Cobas 6000 system with a turbidimetric immune inhibition assay on hemolyzed whole blood or packed red cells. The day-to-day coefficient of variation for HbA1c was 1.9% at 4.4% HbA1c and 1.5% at 10.6% HbA1c.

Fasting lipids, including total cholesterol, high-density lipoprotein cholesterol, and triglycerides, were measured at baseline and follow-up by Quest Diagnostics using standardized procedures. Lipoprotein particle measures were assessed by nuclear magnetic resonance spectroscopy using the LipoProfile-4 platform at Labcorp, Raleigh, North Carolina.

**Glycemic and metabolic trait definitions in VITAL CTSC**

We selected four primary glycemic traits to capture distinct pathways related to type 2 diabetes pathophysiology: HbA1c, insulin sensitivity, 2-hour glucose, and beta-cell function.

HbA1c was used to reflect overall glycemic burden. The oral glucose tolerance test-derived insulin sensitivity index, estimated ISI0,120, was used to assess whole-body insulin sensitivity and was calculated as: ISI0,120 = 0.222 - 0.00333 x BMI - 0.0000779 x insulin120 - 0.000422 x age, where insulin120 represents 2-hour insulin in pmol/L.

Two-hour glucose from the oral glucose tolerance test was used to reflect integrated post-load glucose response. HOMA-B was used to estimate beta-cell function and was calculated as: HOMA-B = 20 x fasting insulin / (fasting glucose - 3.5), where fasting insulin was measured in mU/L and fasting glucose in mmol/L.

In secondary analyses, we evaluated additional glycemic and metabolic traits to aid interpretation, including fasting glucose, 30-minute glucose, fasting insulin, 30-minute insulin, 2-hour insulin, metabolic clearance rate of glucose, HOMA-IR, LPIR score, and diabetes risk index.

The metabolic clearance rate of glucose was calculated as: MCRG = 19.240 - 0.281 x BMI - 0.00498 x insulin120 - 0.333 x glucose120, where insulin120 is 2-hour insulin in pmol/L and glucose120 is 2-hour glucose in mmol/L.

HOMA-IR was calculated as: HOMA-IR = fasting glucose x fasting insulin / 22.5, where fasting glucose was measured in mmol/L and fasting insulin in mU/L.

The lipoprotein insulin resistance score was based on average particle sizes of VLDL, LDL, and HDL and concentrations of large VLDL, small LDL, and large HDL particles. The diabetes risk index was based on lipoprotein and branched-chain amino acid measures and was previously developed to predict incident type 2 diabetes independently of glycemic status.

**Anthropometric and body composition traits in VITAL CTSC**

Body weight, height, waist circumference, and hip circumference were measured at baseline and year 2 using standard clinical equipment by trained personnel. Body mass index was calculated as weight in kilograms divided by height in meters squared. Waist-to-hip ratio was calculated as waist circumference divided by hip circumference.

Body composition was quantified at both time points using dual-energy X-ray absorptiometry on a Discovery W scanner with APEX Software Version 4.2 at the Brigham and Women's Hospital Bone Density Unit of the Skeletal Health and Osteoporosis Center. Measures derived from DXA included total body fat percentage, total fat mass, total lean mass, fat-to-lean mass ratio, truncal fat mass, visceral adipose tissue mass, and limb fat mass.

We selected three primary adiposity traits: fat mass index, truncal fat mass, and visceral adipose tissue mass. Fat mass index was calculated as total fat mass in kilograms divided by height in meters squared and was selected to capture overall adiposity more accurately than BMI alone. Truncal fat mass was selected to capture central adiposity. Visceral adipose tissue mass was selected to capture ectopic fat deposition, which has been independently associated with type 2 diabetes risk.

Visceral adipose tissue was estimated over a 5 cm region at the level of the fourth lumbar vertebra. Subcutaneous adipose tissue, measured from the skin line to the outer abdominal muscle wall, was subtracted from total abdominal adipose tissue, measured within the abdominal muscle walls, to estimate visceral adipose tissue. Additional anthropometric and body composition traits, including BMI, waist-to-hip ratio, fat-to-lean mass ratio, and other DXA-derived measures, were evaluated in secondary analyses.

**UK Biobank biomarker outcomes**

In complementary UK Biobank biomarker analyses, insulin resistance was approximated using the triglyceride-to-HDL cholesterol ratio, calculated from triglycerides, field 30870, and HDL cholesterol, field 30760. Visceral adiposity was quantified using DXA-derived visceral adipose tissue mass, field 23288. Both triglyceride-to-HDL cholesterol ratio and visceral adipose tissue mass were inverse-normal transformed before analysis to approximate normal distributions.

**Incident type 2 diabetes definition in UK Biobank**

Incident type 2 diabetes was identified using linked hospital inpatient records and death registry data. Participants were classified as having incident type 2 diabetes if they had a recorded diagnosis of type 2 diabetes using ICD-10 codes E11.0-E11.9 in field 41270 after the baseline assessment, or if type 2 diabetes was listed as a cause of death in field 40001.

For cases identified through hospital inpatient records, the date of type 2 diabetes onset was defined as the corresponding diagnosis date from field 41280. For participants with type 2 diabetes listed as the underlying cause of death but without a previous hospital diagnosis, the date of death from field 40007 was used as the type 2 diabetes event date.

Time-to-event was calculated from the baseline assessment date, field 53 at instance 0, to the earliest of type 2 diabetes diagnosis, death, loss to follow-up, or end of follow-up on May 31, 2022. Date of loss to follow-up was obtained from field 191.

**Genotyping and pathway-specific polygenic scores in UK Biobank**

UK Biobank participants were genotyped using the UK Biobank Axiom and UK BiLEVE Axiom arrays. Genetic data were imputed using the TOPMed reference panel, with quality control and processing performed as described previously.

We used previously published models to calculate 12 pathway-specific polygenic scores for type 2 diabetes. These scores were developed to estimate genetic susceptibility to type 2 diabetes through distinct pathophysiological pathways. Briefly, the scores were constructed as weighted sums of type 2 diabetes-associated genetic variants, with weights derived from their associations with 110 type 2 diabetes-related quantitative traits using a clustering algorithm. The 12 pathway-specific polygenic scores were standardized to mean 0 and standard deviation 1 before analysis.

We preferentially used directly genotyped variants. When directly genotyped variants were unavailable, imputed variants were used. If a variant was unavailable or ambiguous after imputation, we used proxy variants with linkage disequilibrium r2 >0.8 when available.

**Caffeine metabolism genetic scores**

To account for interindividual genetic variation in caffeine metabolism, we derived caffeine metabolism genetic scores using variants previously associated with circulating caffeine metabolite levels in genome-wide association studies. CMSG2 included two variants with the largest reported effect sizes: rs2472297 near CYP1A2 and rs6968554 near AHR. CMSG4 additionally included rs17685 in POR and rs56113850 near CYP2A6.

Weighted caffeine metabolism genetic scores were calculated by summing the number of effect alleles at each variant multiplied by the corresponding published beta-coefficient. These weighted scores were included as covariates in UK Biobank models.

**Covariate assessment and coding**

In VITAL CTSC, participants self-reported age, sex, race, smoking status, alcohol use, family history of diabetes, and lifestyle factors. Smoking status was modeled as never versus ever smoking. Alcohol use was categorized as daily, weekly, monthly, or rarely/never. Race was modeled as White versus Other because of the limited number of participants in individual non-White racial groups. Physical activity, including exercise and stair climbing, was assessed by questionnaire and quantified as total metabolic equivalent hours per week.

Dietary covariates were assessed using the food frequency questionnaire. Total juice intake and red/processed meat intake were modeled in servings per day. For year 2 analyses, values were estimated by averaging baseline and year 4.5 dietary assessments, consistent with the approach used for coffee intake. BMI was modeled as a continuous covariate in secondary models.

In UK Biobank, baseline covariates were obtained from the first assessment visit. Age and sex were assessed at baseline. Smoking was modeled using smoking status and smoking intensity, including smoking frequency and number of cigarettes per day for current or former smokers, based on fields 3456 and 2887. Alcohol consumption was modeled using self-reported frequency of alcohol intake. Tea intake was assessed as cups per day using field 1488. Physical activity was quantified as total MET-minutes per week for all activities using field 22040.

Models involving pathway-specific polygenic scores were additionally adjusted for genotype measurement batch, UK Biobank assessment center, and the top 10 genetic principal components to account for genetic ancestry and technical variation.

**Statistical analysis in VITAL CTSC**

Clinical and anthropometric traits with skewed distributions were log-transformed before analysis. These included HOMA-B, 2-hour glucose, fat mass index, truncal fat mass, and visceral adipose tissue mass. Distributions of all biomarkers and traits were examined before modeling. For the seven pre-selected primary traits, extreme values were defined as observations beyond +/-5 standard deviations from the mean and were winsorized at the +/-5 standard deviation threshold to reduce the influence of outliers while preserving the analytic sample.

All primary biomarkers and traits were standardized to mean 0 and standard deviation 1 before modeling. Therefore, association estimates represent standard deviation differences in the outcome per one additional cup per day of coffee intake.

Associations between coffee intake and repeated glycemic and adiposity traits were estimated using linear mixed models with participant-specific random intercepts to account for within-person correlation across baseline and year 2 measures. Models were adjusted for age, sex, family history of diabetes, smoking status, race, alcohol use, physical activity, total juice intake, and red/processed meat intake. A second model further adjusted for BMI as a continuous covariate.

Potential effect modification was evaluated using stratified analyses by age, BMI, physical activity, smoking status, sex, and family history of diabetes. Age was categorized as <65 versus >=65 years. BMI was categorized as <25, 25-30, and >30 kg/m2. Physical activity was categorized as <21 versus >=21 MET-hours/week. Smoking status was categorized as never versus former/current smoking.

Seven traits were pre-specified as primary outcomes: HbA1c, insulin sensitivity index, 2-hour glucose, HOMA-B, fat mass index, truncal fat mass, and visceral adipose tissue mass. For these primary traits, a Bonferroni-corrected P value threshold of 0.00714, corresponding to 0.05 divided by 7, was considered statistically significant. Secondary biomarkers and traits were evaluated to aid biological interpretation, and P <0.05 was considered nominally significant.

**Statistical analysis in UK Biobank**

Pathway-specific polygenic scores were standardized to mean 0 and standard deviation 1 before analysis. Cox proportional hazards regression models were used to estimate hazard ratios and 95% confidence intervals for incident type 2 diabetes associated with coffee intake. Coffee intake was modeled both continuously in cups per day and categorically as none, <=1 cup/day, 2-3 cups/day, 4-5 cups/day, and >=6 cups/day.

Time-to-event was calculated from baseline assessment to type 2 diabetes diagnosis, death, loss to follow-up, or the end of follow-up on May 31, 2022, whichever occurred first. Models were adjusted for age, sex, smoking frequency, number of cigarettes per day among current or former smokers, alcohol consumption, tea intake, physical activity, and caffeine metabolism genetic scores. A second model additionally adjusted for BMI.

Non-linear dose-response associations between coffee intake and type 2 diabetes risk were evaluated using Cox proportional hazards models with restricted cubic splines. For spline analyses, the continuous exposure variable was defined as the median coffee intake in cups per day within 20 equally sized quantiles.

We examined associations between each pathway-specific polygenic score and incident type 2 diabetes across categories of coffee intake using Cox regression. These models were adjusted for the covariates listed above and additionally included genotype measurement batch, UK Biobank assessment center, and the top 10 genetic principal components. Multiplicative gene-diet interactions were tested by including an interaction term between continuous coffee intake in cups per day and each pathway-specific polygenic score. Likelihood ratio tests were used to compare models with and without the interaction term. For the 12 pathway-specific polygenic scores, a Bonferroni-corrected interaction threshold of P-interaction <0.00417, corresponding to 0.05 divided by 12, was considered statistically significant.

In complementary UK Biobank biomarker analyses, associations of coffee intake with triglyceride-to-HDL cholesterol ratio and visceral adipose tissue mass were evaluated using linear regression. Both outcomes were inverse-normal transformed before analysis. Models included the same lifestyle and demographic covariates used in the incident type 2 diabetes analyses, with BMI added in a secondary model.

All statistical tests were two-sided. Analyses were conducted using R version 4.3.
